# Total-Body Multiparametric PET Quantification of ^18^F-FDG Delivery and Metabolism in the Study of COVID-19 Recovery

**DOI:** 10.1101/2023.03.26.23287673

**Authors:** Yiran Wang, Lorenzo Nardo, Benjamin A. Spencer, Yasser G. Abdelhafez, Elizabeth J. Li, Negar Omidvari, Abhijit J. Chaudhari, Ramsey D. Badawi, Terry Jones, Simon R. Cherry, Guobao Wang

## Abstract

Conventional whole-body ^18^F-FDG PET imaging provides a semi-quantitative evaluation of overall glucose metabolism without gaining insight into the specific transport and metabolic steps. Here we demonstrate the ability of total-body multiparametric ^18^F-FDG PET to quantitatively evaluate glucose metabolism using macroparametric quantification and assess specific glucose delivery and phosphorylation processes using microparametric quantification for studying recovery from coronavirus disease 2019 (COVID-19).

**Methods:** The study included thirteen healthy subjects and twelve recovering COVID-19 subjects within eight weeks of confirmed diagnosis. Each subject had a dynamic ^18^F-FDG scan on the uEXPLORER total-body PET/CT system for one hour. Semiquantitative standardized uptake value (SUV) and SUV ratio relative to blood (SUVR) were calculated for regions of interest (ROIs) in different organs to measure glucose utilization. Tracer kinetic modeling was performed to quantify microparametric rate constants *K*_1_ and *k*_3_ that characterize ^18^F-FDG blood-to-tissue delivery and intracellular phosphorylation, respectively, and a macroparameter *K*_i_ that represents ^18^F-FDG net influx rate. Statistical tests were performed to examine differences between the healthy controls and recovering COVID-19 subjects. Impact of COVID-19 vaccination was investigated. We further generated parametric images to confirm the ROI-based analysis.

**Results:** We detected no significant difference in lung SUV but significantly higher lung SUVR and *K*_i_ in the recovering COVID-19 subjects, indicating an improved sensitivity of kinetic quantification for detecting the difference in glucose metabolism. A significant difference was also observed in the lungs with the phosphorylation rate *k*_3_, but not with the delivery rate *K*_1_, which suggests it is glucose phosphorylation, not glucose delivery, that drives the observed difference of glucose metabolism in the lungs. Meanwhile, there was no or little difference in bone marrow metabolism measured with SUV, SUVR and *K*_i_, but a significant increase in bone-marrow ^18^F-FDG delivery rate *K*_1_ in the COVID-19 group (*p* < 0.05), revealing a difference of glucose delivery in this immune-related organ. The observed differences were lower or similar in vaccinated COVID-19 subjects as compared to unvaccinated ones. The organ ROI-based findings were further supported by parametric images.

**Conclusions:** Higher lung glucose metabolism and bone-marrow glucose delivery were observed with total-body multiparametric ^18^F-FDG PET in recovering COVID-19 subjects as compared to healthy subjects, which suggests continued inflammation due to COVID-19 during the early stages of recovery. Total-body multiparametric PET of ^18^F-FDG delivery and metabolism can provide a more sensitive tool and more insights than conventional static whole-body ^18^F-FDG imaging to evaluate metabolic changes in systemic diseases such as COVID-19.

## INTRODUCTION

Positron emission tomography (PET) with the radiotracer ^18^F-fluorodeoxyglucose (^18^F-FDG) is a non-invasive *in vivo* molecular imaging technique that reflects glucose metabolism. Conventional whole-body static ^18^F-FDG PET imaging can provide an overall evaluation of glucose utilization throughout the body, but it mixes the specific glucose transport and metabolic steps. Identification and quantification of these specific processes separately require a fast dynamic scanning protocol, which is however limited to a single organ or a confined region by a conventional short axial field-of-view PET scanner. The advent of total-body PET/CT systems such as uEXPLORER (*1*) and other long-axial field-of-view PET scanners (*2,3*) has brought new opportunities for total-body dynamic PET imaging with increased detection sensitivity and simultaneous dynamic imaging of multiple organs (*4*). Combined with tracer kinetic modeling (*5*), total-body dynamic ^18^F-FDG-PET enables a multiparametric quantification method (*6*) that allows quantitative measurement of not only overall glucose utilization, but also the microparametric rates of glucose delivery and phosphorylation (*7*) over the entire body.

Though mostly used in oncology, ^18^F-FDG PET also has the potential for characterizing inflammatory diseases such as vasculitis (*8*), hepatitis (*9*), osteomyelitis (*10*), and recently Coronavirus Disease 2019 (COVID-19) (*11*). COVID-19 primarily attacks the respiratory system, leading to conditions varying from mild manifestations to high-mortality acute symptoms (*12*). Meanwhile, it can affect multiple organs associated with different body systems, including the nervous (*13*), cardiovascular (*14*), and immune systems (*15*). In addition, a variety of prolonged effects of COVID-19 have been reported (*16*–*19*). However, investigations of the whole-body consequences and prolonged effects from COVID-19 are limited, partially due to the lack of an approach for in-depth total-body evaluation.

In this paper, we conducted a quantitative evaluation of glucose utilization in multiple organs of healthy and recovering COVID-19 subjects using total-body multiparametric ^18^F-FDG PET imaging. We analyzed the overall glucose metabolism, and more subtly, the blood-to-tissue glucose delivery and glucose phosphorylation to gain further insight into the metabolic differences induced by COVID-19.

## MATERIALS AND METHODS

### Study Participants and Data Acquisition

With Institutional Review Board approval and written informed consent at the University of California Davis Health, the study includes a cohort of thirteen healthy subjects (all scanned prior to the onset of the COVID-19 pandemic) and twelve COVID-19 subjects. These COVID-19 subjects had mild to moderate symptoms and none of them was hospitalized. Each subject had a total-body one-hour ^18^F-FDG dynamic scan on the 2-meter long uEXPLORER PET/CT system (*20,21*). The PET/CT scans for the COVID-19 subjects were performed within eight weeks of confirmed diagnosis. All COVID-19 subjects tested negative for COVID-19 least two weeks prior to the PET scan. The subjects were injected with 333 ± 45 MBq ^18^F-FDG intravenously immediately after initiating list-mode data acquisition. A total-body ultralow-dose CT scan (140 kVp, 5 mAs) was performed before the PET scan for attenuation correction. Dynamic PET data were reconstructed into 29 frames (6 × 10 s,2 × 30 s,6 × 60 s,5 × 120 s,4 × 180 s,6 × 300 s) with a voxel size of 4 × 4 × 4 mm^3^ using the vendor-provided ordered subset expectation maximization algorithm with four iterations and 20 subsets (*20*).

### Total-body Kinetic Modeling

Regions of interest (ROIs) were placed in various organs and tissues (e.g., brain, liver, lungs, spleen, bone marrow) throughout the entire body on the dynamic images of each subject (see details of ROI placement in Supplemental Table 1). Time-activity curves (TACs) were then extracted from the organ ROIs. In addition, ROI placement and TAC extraction were also done for the ascending aorta (AA) and right ventricle (RV) to acquire image-derived input functions (IDIFs).

A two-tissue irreversible (2Ti) compartmental model was used for modeling the dynamic ^18^F-FDG data (*6*) (Supplemental Fig. 1) following a set of differential equations:

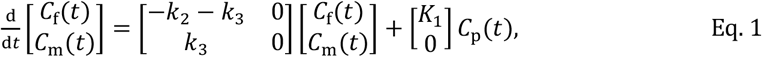

where *C*_p_(*t*), *C*_f_(*t*), and *C*_m_(*t*) are the ^18^F-FDG concentrations in the blood plasma, tissue free-state ^18^F-FDG, and tissue phosphorylated ^18^F-FDG, respectively. *K*_1_ (mL/min/cm^3^) and *k*_2_ (min^-1^) represent the blood-to-tissue and the tissue-to-blood ^18^F-FDG delivery rate, respectively; *k*_3_ (min^-1^) is the ^18^F-FDG phosphorylation rate. This irreversible model assumes negligible ^18^F-FDG dephosphorylation, i.e., the ^18^F-FDG dephosphorylation rate *k*_4_ (min^-1^) is zero.

The activity that can be directly measured with PET is modeled as a combination of the concentrations in blood and tissue,

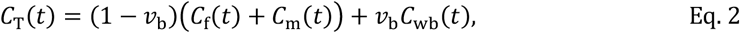

where *v*_b_ is the blood volume fraction and *C*_wb_(*t*) is the ^18^F-FDG concentration in the whole blood that is approximated by *C*_p_(*t*). A time delay correction was applied to account for the time difference from where the IDIF is extracted to the arrival in a tissue (*6*), and different image-derived input functions were used as appropriate for the kinetic modeling of different organs. The IDIF for most organs is the ascending aorta TAC:

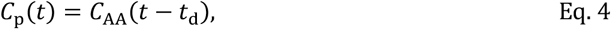

except for the lungs for which the IDIF is the right ventricle TAC:

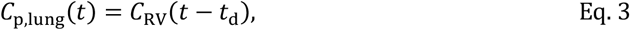

where *t*_d_ (s) is the time delay correction parameter.

All the kinetic parameters including the time delay were jointly estimated through a non-linear least-square fitting method (*6*) with a weighting factor that considers the time length of each frame and nuclear decay (*22*).

### Macroparametric and Microparametric Quantification

The macro-parameter *K*_i_, denoting ^18^F-FDG net influx rate, is commonly used to characterize overall glucose metabolism and is calculated by:

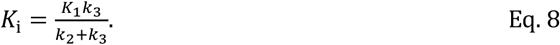

We also applied semi-quantitative standardized uptake value (SUV) (*23*) and SUV ratio relative to blood (SUVR) (*24*) using the last dynamic frame (55-60 min) to evaluate the overall glucose metabolism. RV was used for the lung SUVR calculation, and AA was used for the SUVR calculation of all other organs.

In addition to the measures of overall glucose metabolism by SUV, SUVR, and *K*_i_, we also used the microparameters of the 2Ti kinetic model, specifically the ^18^F-FDG delivery rate *K*_1_ and phosphorylation rate *k*_3_, to gain insight into the individual molecular processes of glucose utilization. The ability of this microparametric quantification is a feature that distinguishes compartmental modeling from whole-body static imaging or whole-body dynamic imaging with a simplified graphical analysis method (e.g., the Patlak plot).

### Statistical Analysis

Statistical analysis in this study was performed using an unpaired, two-tailed T test and the Mann-Whitney U test on SUV, SUVR and parametric PET metrics to investigate metabolic differences in the recovering COVID-19 subjects compared to the healthy controls. In addition, the tests were performed on lung CT ROIs (Supplemental Table 1) for complementary information. Effect of vaccination was also investigated when appropriate using the statistical tests between the vaccinated and the unvaccinated COVID-19 recovering groups to study the potential influence of vaccination (*25,26*). All statistical data analyses were conducted using MATLAB (Mathworks, MA). P-values of less than 0.05 were considered statistically significant.

For organs that demonstrated a difference in the glucose metabolism, the Pearson correlation analysis between *K*_i_ and micro-parameters *K*_1_, *k*_2_, and *k*_3_ was also conducted to further understand their association.

### Parametric Imaging of COVID-19

In addition to the ROI-based analysis, voxel-wise parametric images were generated for the healthy and the recovering COVID-19 subjects using the 2Ti compartmental model. Kernel smoothing was applied to both the dynamic images and parametric images for noise reduction (*6*). To make the comparison of parametric images more focused on organs of interest, masking was used to visualize individual organs or tissues (e.g., lung or bone marrow) within the parametric images for inter-subject comparisons.

## RESULTS

### Patient Characteristics

A summary of patient characteristics is provided in Supplemental Table 2. The healthy subjects include six males and seven females with age 49 ± 15 y and weight 82 ± 18 kg. The COVID-19 subjects include three males, nine females with age 41 ± 10 y and weight 84 ± 25 kg. There was no statistical difference between the two groups in age, weight, body mass index (BMI), blood glucose level, or fasting time before the PET scan using the unpaired T test and the U test. In addition, there was no statistical difference in lung CT values and in the SUV of the input function (neither AA nor RV) between the two groups.

### Dynamic Images and TACs

Total-body dynamic ^18^F-FDG PET images of a representative healthy subject and a recovering COVID-19 subject are shown in Fig. 1A. Fig. 1B shows four examples of the TACs in the form of SUV and SUVR over time. The most notable finding was the increased lung SUVR in the recovering COVID-19 group compared to the healthy group, while the bone marrow SUVR and spleen SUVR of recovering COVID-19 group also tended to be higher.

**FIGURE 1.**
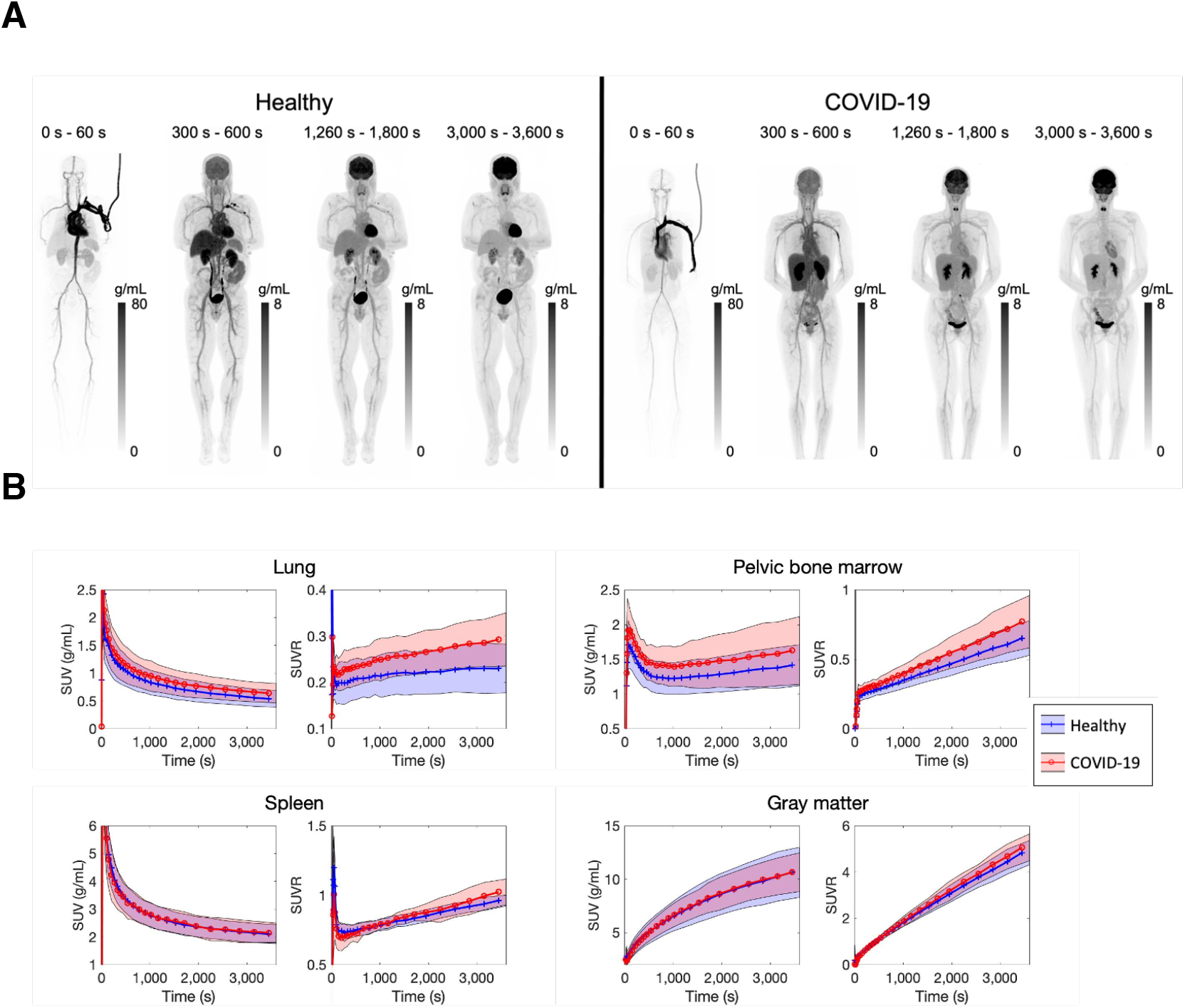
(A) Total-body dynamic ^18^F-FDG PET images of a healthy subject and a recovering COVID-19 subject. Shown are maximum intensity projections. (B) Averaged TACs (shown as SUV and SUVR) of four organs of interest (lung, pelvic bone marrow, spleen, and gray matter) of the thirteen healthy and the twelve recovering COVID-19 subjects. The averaged values are shown as the solid lines, and the standard deviations are shown as the bands.

### Comparison of Overall Glucose Utilization in Multiple Organs

Table 1 summarizes the SUV, SUVR, and *K*_i_ of the healthy and the recovering COVID-19 groups along with group comparison results for 11 different organ ROIs. There was no significant difference in lung SUV between the two groups (*p* > 0.1) (Fig. 2). However, there was a statistically significant increase of ∼120% in lung *K*_i_ in the COVID-19 group (*p* ≈ 0.01). SUVR showed a difference (∼25% increase) but to a lower degree.

**TABLE 1.** Comparison of the ^18^F-FDG metabolic metrics SUV, SUVR, and *K*_i_ between the healthy subjects and recovering COVID-19 subjects in multiple organs/tissues.

| Organ/tissue | Metric | Healthy group<br>(mean ± sd) | COVID-19 recovering<br>group<br>(mean ± sd) | $p_T$ | $p_U$ |
| --- | --- | --- | --- | --- | --- |
| Lung | SUV (g/mL) | 0.54±0.16 | 0.64±0.18 | 0.15 | 0.22 |
| | SUVR | 0.230±0.055 | 0.293±0.060 | 0.012 | 0.018 |
| | $K_i$ (mL/min/cm<sup>3</sup>) | 0.00038±0.00033 | 0.00084±0.00045 | 0.0075 | 0.011 |
| Myocardium | SUV (g/mL) | 7.5±3.5 | 5.8±2.8 | 0.21 | 0.20 |
| | SUVR | 3.4±1.6 | 2.8±1.4 | 0.38 | 0.34 |
| | $K_i$ (mL/min/cm<sup>3</sup>) | 0.055±0.033 | 0.043±0.025 | 0.31 | 0.37 |
| Liver | SUV (g/mL) | 2.64±0.44 | 2.56±0.40 | 0.65 | 0.61 |
| | SUVR | 1.208±0.060 | 1.218±0.061 | 0.69 | 0.68 |
| | $K_i$ (mL/min/cm<sup>3</sup>) | 0.00279±0.00094 | 0.00330±0.00086 | 0.17 | 0.17 |
| Spleen | SUV (g/mL) | 2.11±0.35 | 2.15±0.36 | 0.74 | 0.93 |
| | SUVR | 0.963±0.041 | 1.024±0.097 | 0.048 | 0.053 |
| | $K_i$ (mL/min/cm<sup>3</sup>) | 0.0037±0.0010 | 0.0049±0.0018 | 0.055 | 0.087 |
| Spine bone marrow | SUV (g/mL) | 2.06±0.38 | 2.21±0.59 | 0.43 | 0.57 |
| | SUVR | 0.95±0.17 | 1.05±0.21 | 0.21 | 0.22 |
| | $K_i$ (mL/min/cm<sup>3</sup>) | 0.0072±0.0015 | 0.0080±0.0023 | 0.35 | 0.50 |
| Pelvic bone marrow | SUV (g/mL) | 1.42±0.31 | 1.63±0.51 | 0.22 | 0.43 |
| | SUVR | 0.65±0.13 | 0.77±0.20 | 0.087 | 0.13 |
| | $K_i$ (mL/min/cm<sup>3</sup>) | 0.0050±0.0012 | 0.0059±0.0019 | 0.19 | 0.24 |
| Thigh muscle | SUV (g/mL) | 0.57±0.16 | 0.58±0.12 | 0.92 | 0.93 |
| | SUVR | 0.262±0.056 | 0.279±0.065 | 0.50 | 0.72 |
| | $K_i$ (mL/min/cm<sup>3</sup>) | 0.00168±0.00057 | 0.00179±0.00059 | 0.65 | 0.89 |
| Gray matter | SUV (g/mL) | 10.7±2.4 | 10.7±1.9 | 0.99 | 0.76 |
| | SUVR | 4.84±0.54 | 5.07±0.60 | 0.33 | 0.31 |
| | $K_i$ (mL/min/cm<sup>3</sup>) | 0.0476±0.0062 | 0.0487±0.0061 | 0.65 | 0.68 |
| White matter | SUV (g/mL) | 4.5±1.6 | 3.9±1.0 | 0.28 | 0.22 |
| | SUVR | 2.03±0.45 | 1.85±0.31 | 0.26 | 0.46 |
| | $K_i$ (mL/min/cm<sup>3</sup>) | 0.0168±0.0051 | 0.0148±0.0046 | 0.33 | 0.50 |
| Brainstem | SUV (g/mL) | 6.1±1.3 | 5.84±0.82 | 0.55 | 0.68 |
| | SUVR | 2.78±0.24 | 2.79±0.34 | 0.90 | 0.85 |
| | $K_i$ (mL/min/cm<sup>3</sup>) | 0.0247±0.0023 | 0.0241±0.0033 | 0.62 | 0.46 |
| Cerebellum | SUV (g/mL) | 7.3±1.3 | 6.99±0.77 | 0.49 | 0.50 |
| | SUVR | 3.34±0.28 | 3.35±0.27 | 0.93 | 0.89 |
| | $K_i$ (mL/min/cm<sup>3</sup>) | 0.0300±0.0033 | 0.0300±0.0030 | 1.0 | 1.0 |

**FIGURE 2.**
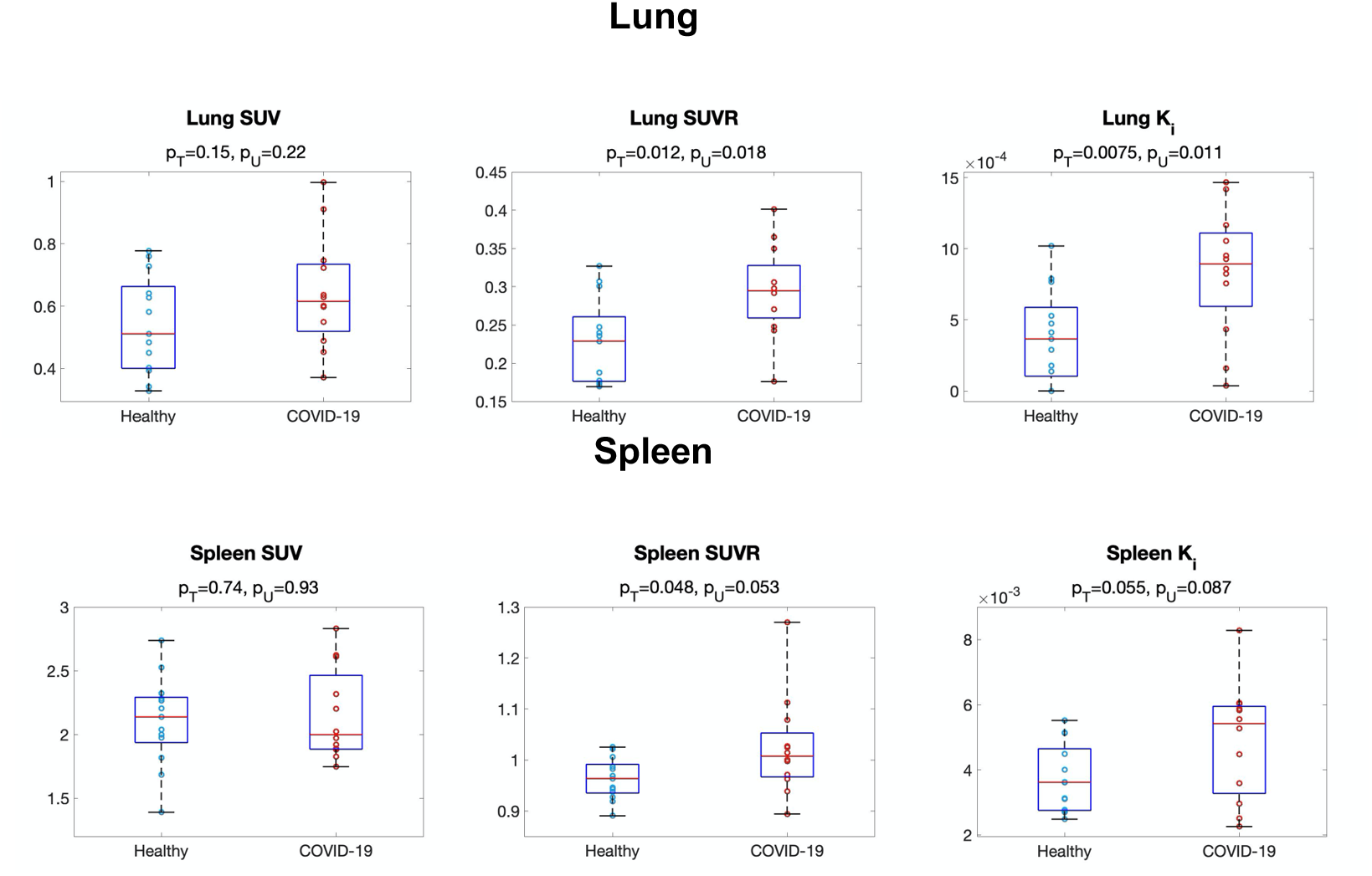
Comparison of ^18^F-FDG glucose metabolism in the lung (top) and spleen (bottom) between the healthy and recovering COVID-19 groups using SUV, SUVR (both at 55 - 60 min), and *K*_i_. *p*_T_ and *p*_U_ are the p-values of the T test and the Mann-Whitney U test, respectively.

Spleen metabolism and bone marrow metabolism also tended to increase, though did not always achieve a statistical significance, as shown in Table 1 and the boxplots in Fig. 2. *K*_i_ produced a larger group difference than SUV, while SUVR was comparable to *K*_i_. We did not observe a statistically significant difference with SUV, SUVR, and *K*_i_ in other organs (e.g., brain, liver). Based on the above analyses, the lung, bone marrow, and spleen were selected for further study of microparametric quantification.

### Microparametric Quantification of the Lungs

Table 2 shows the analysis of microparametric quantification of the lungs. The correlation between each microparameter and lung *K*_i_ is also included using all subject data. Neither *K*_1_ Nor *k*_2_ detected any group difference (*p* > 0.6). *k*_3_ was much higher in the COVID-19 group (*p* < 0.05), as further shown in Fig. 3A. Also, *k*_3_ had the strongest correlation with *K*_i_ (r = 0.56, *p* = 0.0035) among the three microparameters (Fig. 3B), while the correlations of *K*_1_ and *k*_2_ with *K*_i_ were poor (*p* > 0.25). The findings suggested that increased ^18^F-FDG phosphorylation (as quantified by *k*_3_) might be the main driving factor for the increased lung ^18^F-FDG metabolism (assessed by *K*_i_) in COVID-19 recovery.

**TABLE 2.** Healthy *vs*. recovering COVID-19 comparison of lung micro kinetic parameters *K*_1_, *k*_2_, and *k*_3_ and the correlation between the microparameters and lung ^18^F-FDG net influx rate *K*_i_.

| Kinetic parameter | Healthy vs. COVID-19 group comparison | | | | Correlation with $K_i$ | |
| --- | --- | --- | --- | --- | --- | --- |
| | Healthy group<br>(mean ± sd) | COVID-19<br>recovering group<br>(mean ± sd) | $p_T$ | $p_U$ | $r$ | $p$ |
| $K_1$ (mL/min/cm<sup>3</sup>) | 0.018±0.022 | 0.017±0.019 | 0.89 | 0.98 | 0.23 | 0.26 |
| $k_2$ (min<sup>-1</sup>) | 0.32±0.33 | 0.26±0.25 | 0.61 | 0.81 | 0.17 | 0.42 |
| $k_3$ (min<sup>-1</sup>) | 0.0079±0.0071 | 0.021±0.023 | 0.049 | 0.011 | 0.56 | 0.0035 |

**FIGURE 3.**
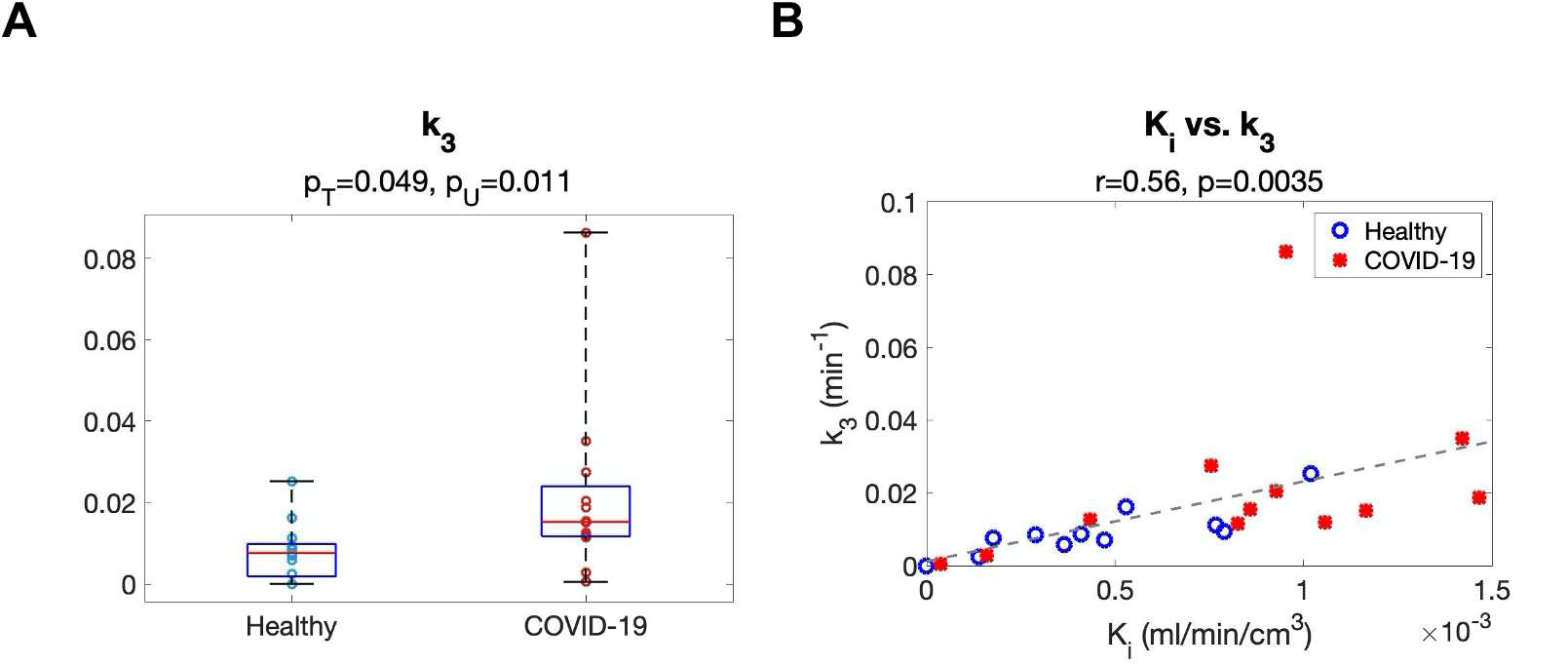
Study of lung kinetic parameters in the healthy and the recovering COVID-19 groups. (A) Comparison of ^18^F-FDG phosphorylation rate *k*_3_ between the two groups. (B) Correlation between *k*_3_ and ^18^F-FDG net influx rate *K*_i_ among the subjects.

### Microparametric Quantification of Bone Marrow

The microparametric quantification results of bone marrow are summarized in Table 3. While bone marrow metabolism did not show a statistical difference between the two groups as measured with SUVR and *K*_i_ (Table 1), bone-marrow ^18^F-FDG delivery rate *K*_1_ was higher in the COVID-19 subjects, as also shown in Fig. 4. The pelvic bone marrow *K*_1_ was ∼20% higher in the COVID-19 group than in the healthy group (*p* < 0.05) (Table 3). In comparison, no statistical significances were observed in *k*_3_. In contrast to the results in the lungs, here the bone marrow microparameters *K*_1_, *k*_2_, and *k*_3_ all had strong correlations with *K*_i_ with *p* < 0.05, though the correlation of *K*_1_ with *K*_i_ remained weaker (Table 3). Similar results were also found in the spine bone marrow.

**TABLE 3.** Healthy *vs*. recovering COVID-19 comparison of bone marrow micro kinetic parameters *K*_1_, *k*_2_, and *k*_3_ and the correlation between the microparameters and bone marrow ^18^F-FDG net influx rate *K*_i_.

| Bone marrow type | Kinetic parameter | Healthy vs. COVID-19 recovering comparison | | | | Correlation with $K_i$ | |
| --- | --- | --- | --- | --- | --- | --- | --- |
| | | Healthy group (mean ± sd) | COVID-19 recovering group (mean ± sd) | $p_T$ | $p_U$ | $r$ | $p$ |
| Spine | $K_1$ (mL/min/cm<sup>3</sup>) | 0.221±0.055 | 0.285±0.089 | 0.041 | 0.068 | 0.46 | 0.020 |
| | $k_2$ (min<sup>-1</sup>) | 0.76±0.19 | 0.92±0.31 | 0.14 | 0.20 | 0.45 | 0.023 |
| | $k_3$ (min<sup>-1</sup>) | 0.0261±0.0061 | 0.027±0.013 | 0.73 | 0.76 | 0.78 | <0.0001 |
| Pelvic | $K_1$ (mL/min/cm<sup>3</sup>) | 0.122±0.026 | 0.149±0.037 | 0.042 | 0.047 | 0.66 | 0.0003 |
| | $k_2$ (min<sup>-1</sup>) | 0.573±0.081 | 0.64±0.14 | 0.17 | 0.26 | 0.51 | 0.0090 |
| | $k_3$ (min<sup>-1</sup>) | 0.0246±0.0060 | 0.0262±0.0088 | 0.61 | 0.81 | 0.85 | <0.0001 |

**FIGURE 4.**
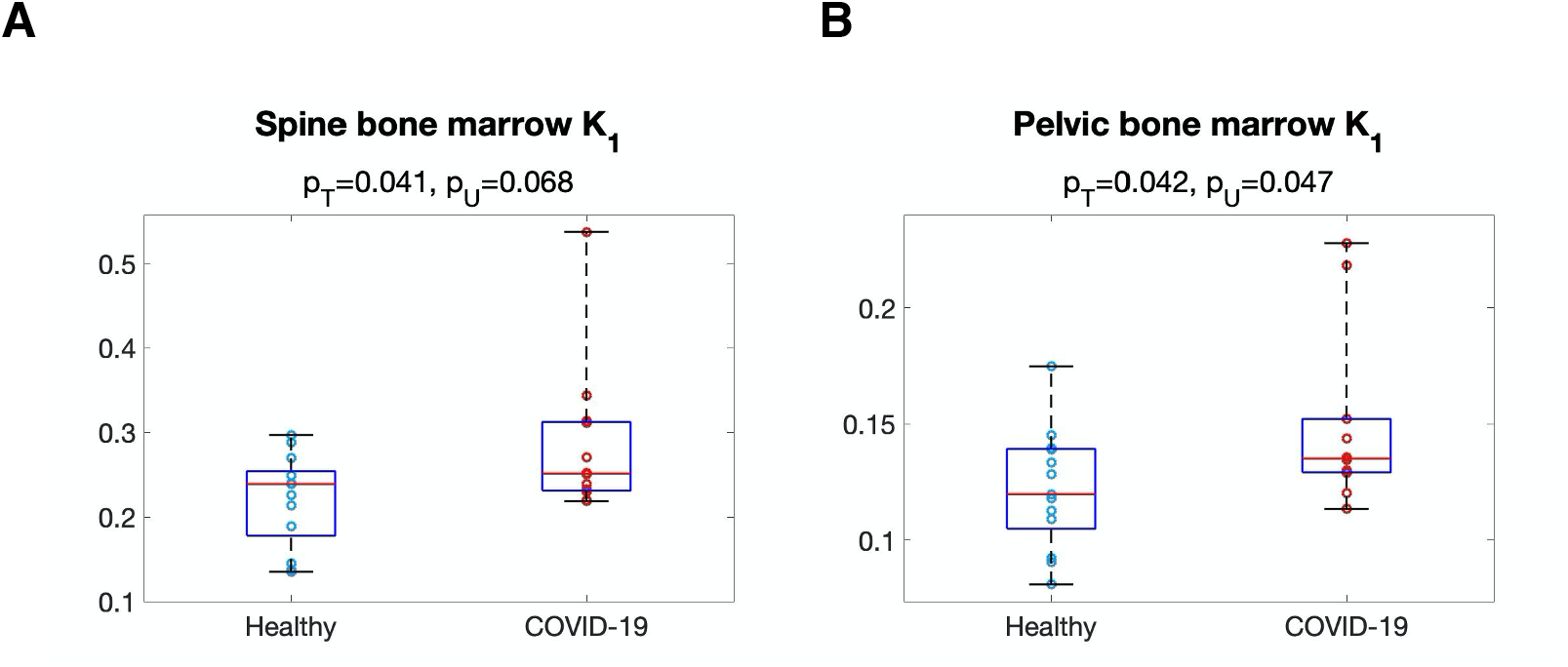
Comparison of ^18^F-FDG delivery rate *K*_1_ of the (A) spine bone marrow and (B) pelvic bone marrow between the healthy and the recovering COVID-19 groups.

### Microparametric Quantification of the Spleen

Table 4 shows the microparametric quantification results for the spleen. *k*_3_ was ∼45% higher in the COVID-19 group (Fig. 5A), while *K*_1_ and *k*_1_ did not show a significant group difference (*p* > 0.3). *k*_3_ correlated the most strongly with *K*_i_ (*r* = 0.98, *p* < 0.0001) among the three microparameters (Fig. 5B), indicating that the increased trend in spleen metabolism (represented by SUVR and *K*_i_) was dominated by the increased ^18^F-FDG phosphorylation. Overall, the observed changes in the spleen were similar to that of the lungs but with a weaker statistical significance.

**TABLE 4.** Healthy *vs*. recovering COVID-19 comparison of spleen microparameters *K*_1_, *k*_2_, and *k*_3_ and the correlation between the microparameters and spleen ^18^F-FDG net influx rate *K*_i_.

| Kinetic parameter | Healthy vs. COVID-19 group comparison | | | | Correlation with $K_i$ | |
| --- | --- | --- | --- | --- | --- | --- |
| | Healthy group<br>(mean ± sd) | COVID-19<br>recovering group<br>(mean ± sd) | $p_T$ | $p_U$ | $r$ | $p$ |
| $K_1$ (mL/min/cm<sup>3</sup>) | 1.61±0.75 | 1.31±0.88 | 0.37 | 0.40 | -0.550 | 0.0044 |
| $k_2$ (min<sup>-1</sup>) | 2.5±1.0 | 2.1±1.2 | 0.34 | 0.40 | -0.426 | 0.034 |
| $k_3$ (min<sup>-1</sup>) | 0.0062±0.0024 | 0.0090±0.0041 | 0.047 | 0.097 | 0.98 | <0.0001 |

**FIGURE 5.**
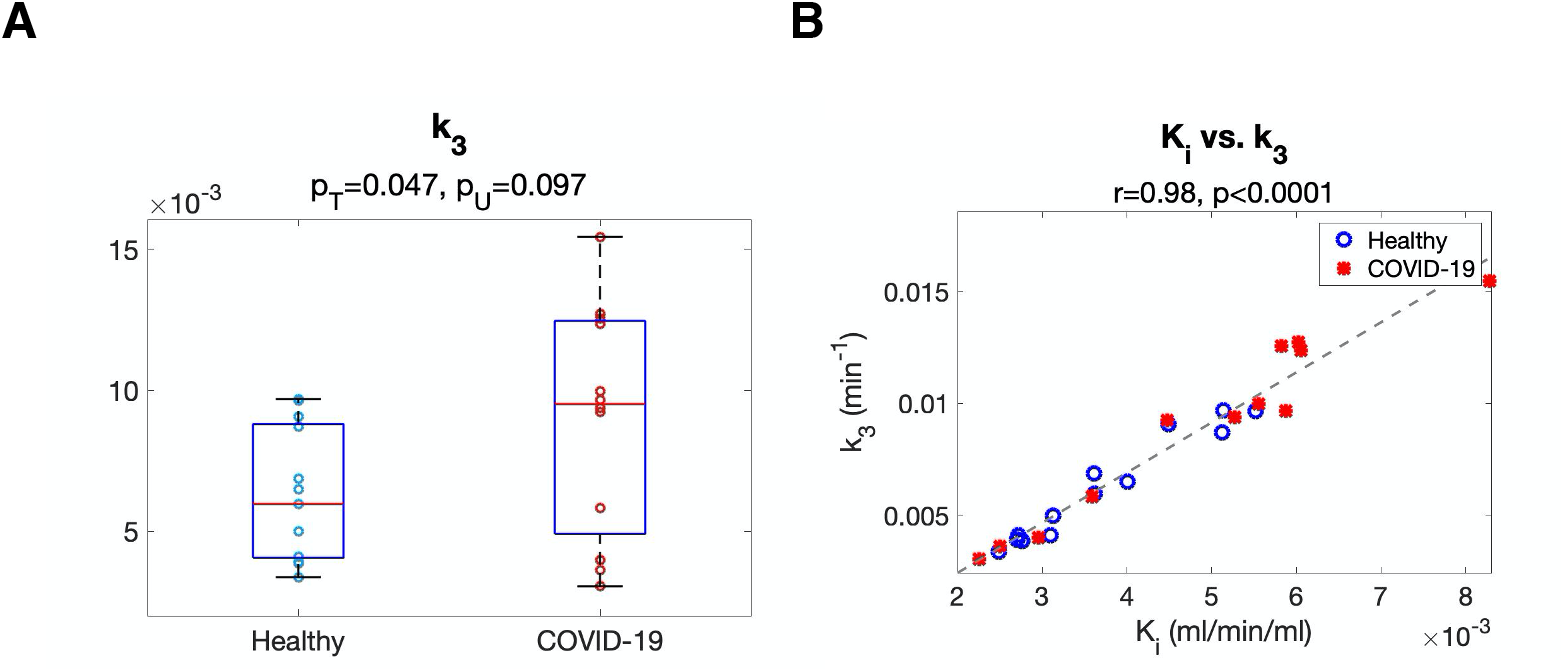
Study of microparametric quantification in the spleen. (A) Comparison of *k*_3_ between the two groups. (B) Correlation between *k*_3_ and *K*_i_ among the subjects.

### Effect of Vaccination

Among the COVID-19 subjects, five were unvaccinated and seven were vaccinated prior to their PET scans (Supplemental Table 2). There was no statistical difference in age, BMI, blood sugar level between the unvaccinated and vaccinated COVID-19 subjects (*p* > 0.2). Lung *K*_i_ was significantly higher in unvaccinated COVID-19 subjects (0.00114±0.00029 mL/min/cm^3^) as compared to healthy subjects (0.00038±0.00033 mL/min/cm^3^) with *p* < 0.001, as shown in Fig. 6A. Lung *K*_i_ was reduced in vaccinated COVID-19 subjects (0.00062±0.00042 mL/min/cm^3^) but still slightly higher than in healthy controls.

**FIGURE 6.**
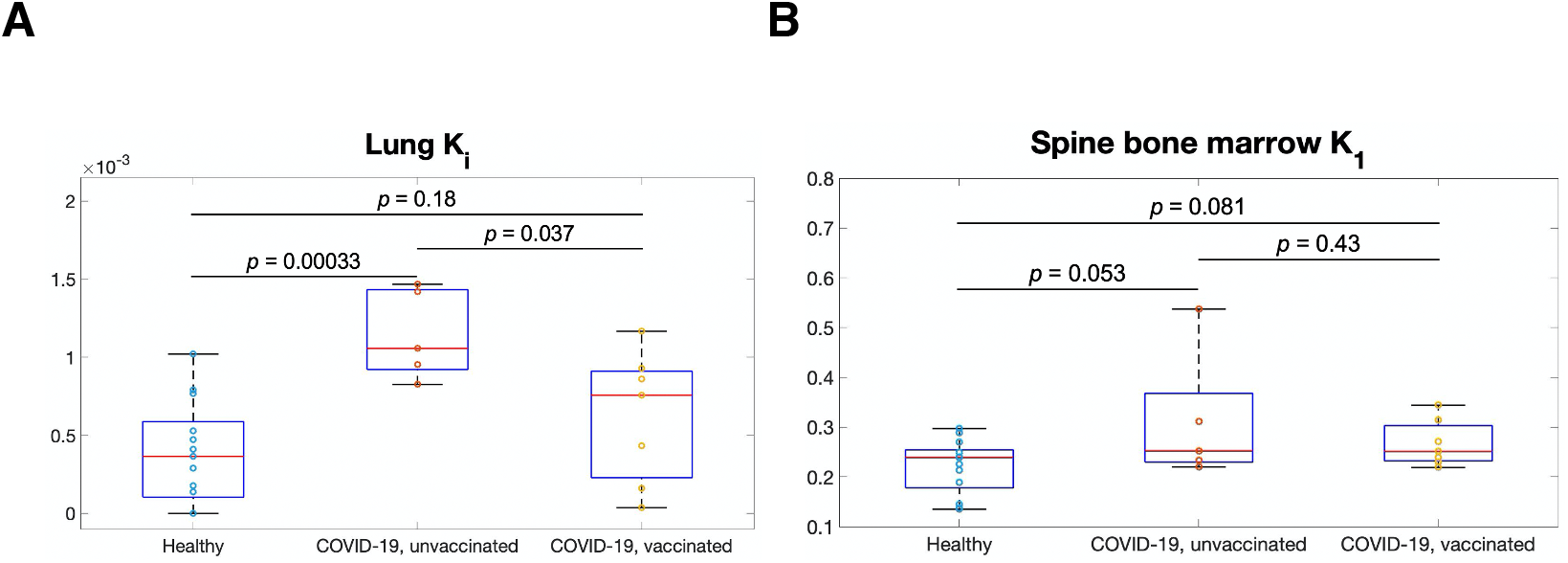
Evaluation of unvaccinated and vaccinated COVID-19 subjects as compared to healthy subjects. (A) Lung ^18^F-FDG net influx rate *K*_i_. (B) Spine bone-marrow ^18^F-FDG delivery rate *K*_1_. P values were calculated using the unpaired T test.

Spine bone-marrow *K*_1_ of both unvaccinated and vaccinated COVID-19 subjects were higher than that of healthy subjects (Fig. 6B). But it did not differ significantly between unvaccinated and vaccinated COVID-19 subjects. No statistical effect of vaccination was noted in other organs of recovering COVID-19 subjects.

### Parametric Imaging of Recovering COVID-19

Fig. 7 shows the parametric images of the lungs and bone marrow from healthy subjects and COVID-19 subjects. The lung images of SUVR, *K*_i_ and *k*_3_ showed enhanced contrast between the healthy and the recovering COVID-19 compared to SUV (Fig. 7A) through visual inspection, supporting the ROI-based analyses. The spine bone marrow (Fig. 7B) and pelvic bone marrow (Supplemental Fig. 2A) images of *K*_i_ and *K*_1_ showed increased contrast between the two subjects than SUV. The SUVR and *K*_i_ images of the spleen also tended to have higher contrast as compared to the SUV images (Supplemental Fig. 2B). These observations are consistent with the ROI-based findings.

**FIGURE 7.**
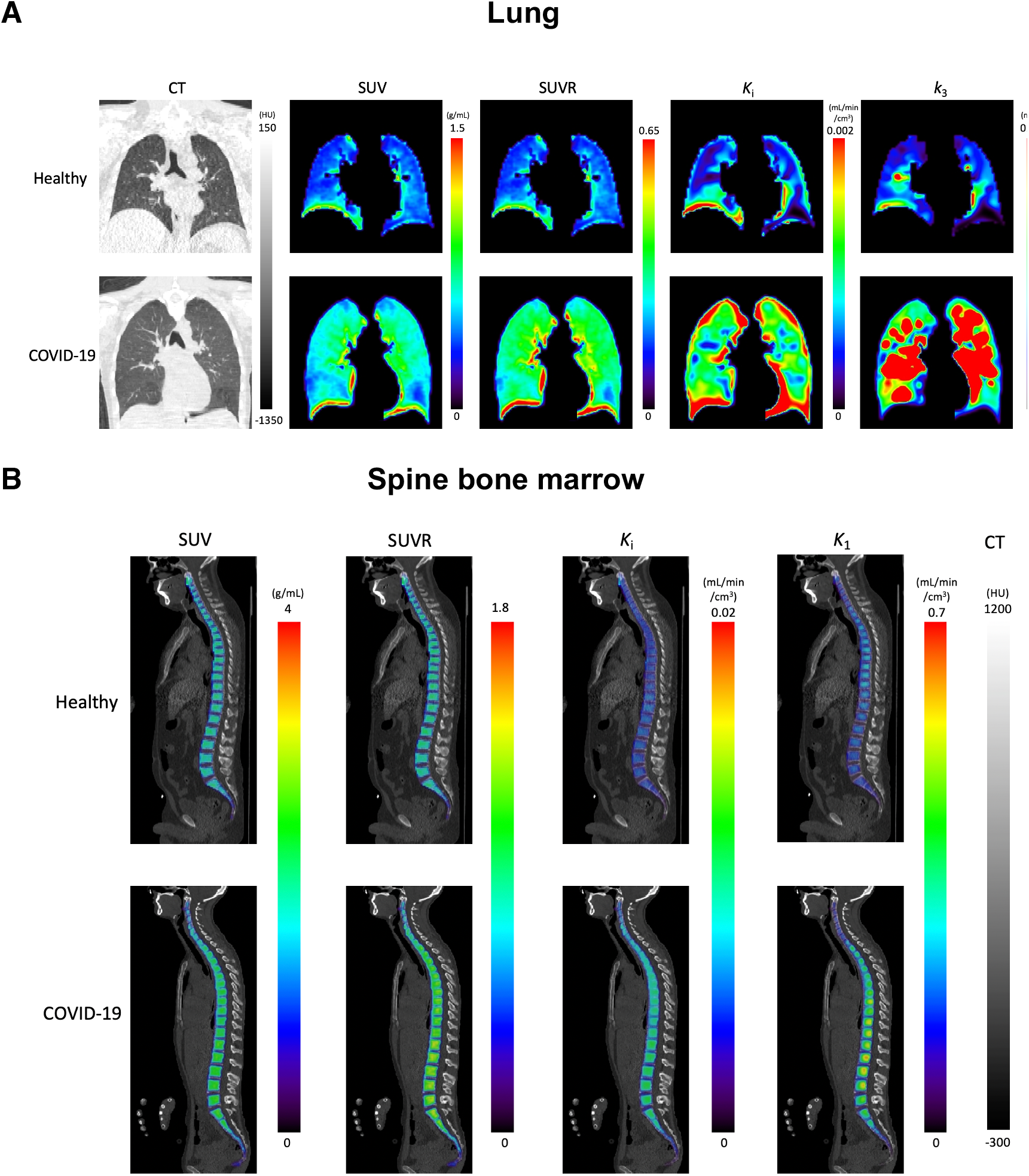
Parametric images of example healthy subjects and COVID-19 subjects. (A) Lung CT, ^18^F-FDG SUV, SUVR, and parametric images of ^18^F-FDG net influx rate *K*_i_, and ^18^F-FDG phosphorylation rate *k*_3_. The coronal slices are selected as the mid of trachea carina. (B) Spine bone marrow images of ^18^F-FDG SUV, SUVR, and parametric images ^18^F-FDG net influx rate *K*_i_, and blood-to-tissue ^18^F-FDG delivery rate *K*_1_. The PET images are masked for the bone marrow region and overlaid on the CT images.

## DISCUSSION

In this study, we evaluated the metabolic differences in multiple organs between recovering COVID-19 subjects and healthy controls using total-body dynamic ^18^F-FDG PET combined with kinetic modeling. We detected increased metabolism using *K*_i_ in the lung, while SUV gave no group differentiation (Table 1 and Fig. 2). The inability of SUV to distinguish the groups is likely due to its semi-quantitative nature and being susceptible to confounding factors (*23*). SUVR showed a larger group difference than SUV but did not outperform *K*_i_. These results suggest the power of kinetic quantification for assessing glucose metabolism. The increased lung metabolism (measured by *K*_i_ and SUVR) in the COVID-19 group may indicate continued inflammation during the early stages of recovery. Previous dynamic lung ^18^F-FDG PET studies have associated increased lung *K*_i_ with pulmonary inflammation in multiple conditions, such as acute lung injury (*27*) and chronic obstructive pulmonary disease (*28*). Meanwhile, prolonged lung inflammation caused by COVID-19 has also been reported, which can last more than 60 days after infection even for asymptomatic and patients with mild cases (*29,30*).

Another advantage of compartmental modeling is microparametric quantification. According to the analysis in the lungs, ^18^F-FDG phosphorylation rate *k*_3_ is the parameter that was responsible for the healthy *vs*. COVID-19 group difference in *K*_i_ (Fig. 3, Fig. 7A) and correlated best with *K*_i_ among different microparameters (Table 2). The result implied that increased ^18^F-FDG phosphorylation may be the main driving factor for increased lung metabolism during the early stages of recovery from COVID-19, while the glucose delivery rate *K*_1_ in the lungs did not differ between COVID-19 and healthy controls (Table 2). These findings are also consistent with previous animal studies that observed *k*_3_ increases in lung inflammation and the association between *K*_i_ and *k*_3_ (*27,31*).

Bone marrow demonstrated a significant change of *K*_1_ in the recovering COVID-19 group as compared to healthy subjects (Fig. 4, Fig. 7B), but no differences were observed with SUV, SUVR or *K*_i_ that reflects the overall ^18^F-FDG metabolism (Table 1). This result further indicates the importance of microparametric quantification. Bone marrow is essential for immunoregulation and is the origin of immune cells (*32*). Animal studies have reported that bone marrow cells play an important role in the repair of the injured lung during lung inflammation (*33,34*). Hence, the increased ^18^F-FDG delivery represented by *K*_1_ may be associated with immune system response during COVID-19 recovery. Given that ^18^F-FDG *K*_1_ of liver was also demonstrated to associate with hepatic inflammation in fatty liver disease (*9,35*), the interplay between *K*_1_ and inflammation reaction, and the potential of *K*_1_ as a biomarker of disease, are worth more studies to explore its clinical applications.

The spleen tended to have higher metabolism in the COVID-19 group, as represented by *K*_i_ or SUVR (Table. 1). This observation is consistent with the splenic ^18^F-FDG uptake increase reported in previous studies of COVID-19 (*36*) and other infectious diseases (*37*). As an immune organ, the spleen plays an important role in the immune response to COVID-19 (*38*) and the immune response may lead to increased metabolism.

Our study also separated the unvaccinated and vaccinated COVID-19 groups to evaluate the potential effect of vaccination (Fig. 6). The lower lung *K*_i_ in the vaccinated group may indicate reduced lung inflammation due to the protection of vaccination. The data also suggest that the observed differences in the lungs and bone marrow between the recovering COVID-19 group and healthy subjects are more likely a direct result of COVID-19, not due to vaccination.

The study has several limitations. The pilot study cohort is relatively small. With an increased sample size, it may be possible to further observe some group differences that were not statistically significant in the current study. Although there is no statistical difference in age, weight, BMI, and blood sugar level between healthy subjects and recovering COVID-19 subjects, the unpaired age and gender can introduce potential risks of biased observation. The study lacks histopathology or clinical laboratory data to elaborate the reason for the differences in ^18^F-FDG kinetics between the two groups. The statistical analysis in this pilot study was not corrected for possible family-wise error rate as the focus of this work is on comparing parametric metrics with SUV. Confirmation of the physiologic findings from this study will require a larger sample size with an appropriate correction for multiple comparisons.

Our next steps are to use a similar methodology to study the impact of long COVID-19 on individual subjects. The interplay and correlation of tracer kinetics among different organs will be also studied in the future. In addition, the results from this pilot work suggest future study designs should focus more on immune-related metabolic changes, e.g., by tracking macrophage (*39*) or neutrophil (*40*) or monitoring serum inflammatory factors.

## CONCLUSIONS

With total-body multiparametric PET, increased lung ^18^F-FDG metabolism (measured by *K*_i_) and increased bone-marrow ^18^F-FDG delivery (measured by *K*_1_) were detected in recovering COVID-19 subjects as compared to healthy subjects. The changes may be associated with continued inflammation and immune response during the early stages of recovery from COVID-19. These findings are otherwise missed or not possible to find if standard SUV measures are used. Total-body multiparametric ^18^F-FDG PET can be a more sensitive tool than conventional whole-body ^18^F-FDG imaging for detecting subtle changes and may be used for studying post-acute sequelae of COVID-19.

## Data Availability

All data produced in the present study are available upon reasonable request to the authors.

## ACKNOWLEDGEMENTS

We gratefully acknowledge the technologists and staff, particularly Lynda E. Painting, of the EXPLORER Molecular Imaging Center (EMIC), for their assistance in patient consent and data acquisition.

## KEY POINTS

### QUESTION

Compared to standard whole-body ^18^F-FDG-PET imaging, is there any benefit from using total-body multiparametric ^18^F-FDG PET for studying COVID-19 recovery?

### PERTINENT FINDINGS

Higher ^18^F-FDG net influx and phosphorylation in the lung and higher ^18^F-FDG blood-to-tissue delivery in bone marrow were detected in recovering COVID-19 subjects compared to healthy subjects, while no statistical difference was detected using SUV.

### IMPLICATIONS FOR PATIENT CARE

Total-body multiparametric ^18^F-FDG PET may offer a more sensitive tool for quantitative assessment of multi-organ effects in COVID-19 recovery than SUV and may be used to study long COVID-19.

---

**SUPPLEMENTAL FIGURE 1.**
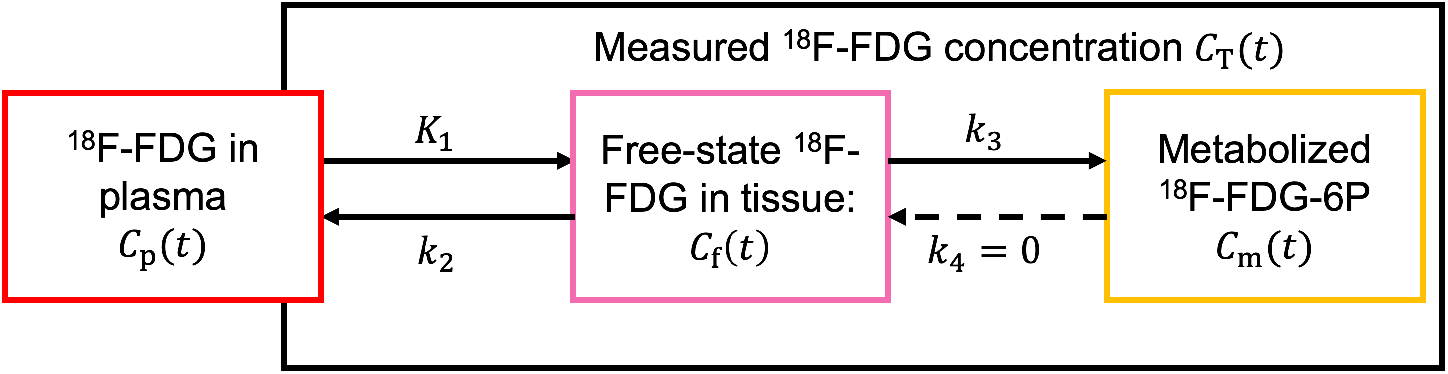
The tracer kinetics of ^18^F-FDG is described by a two-tissue irreversible (2Ti) compartmental model.

**SUPPLEMENTAL FIGURE 2.**
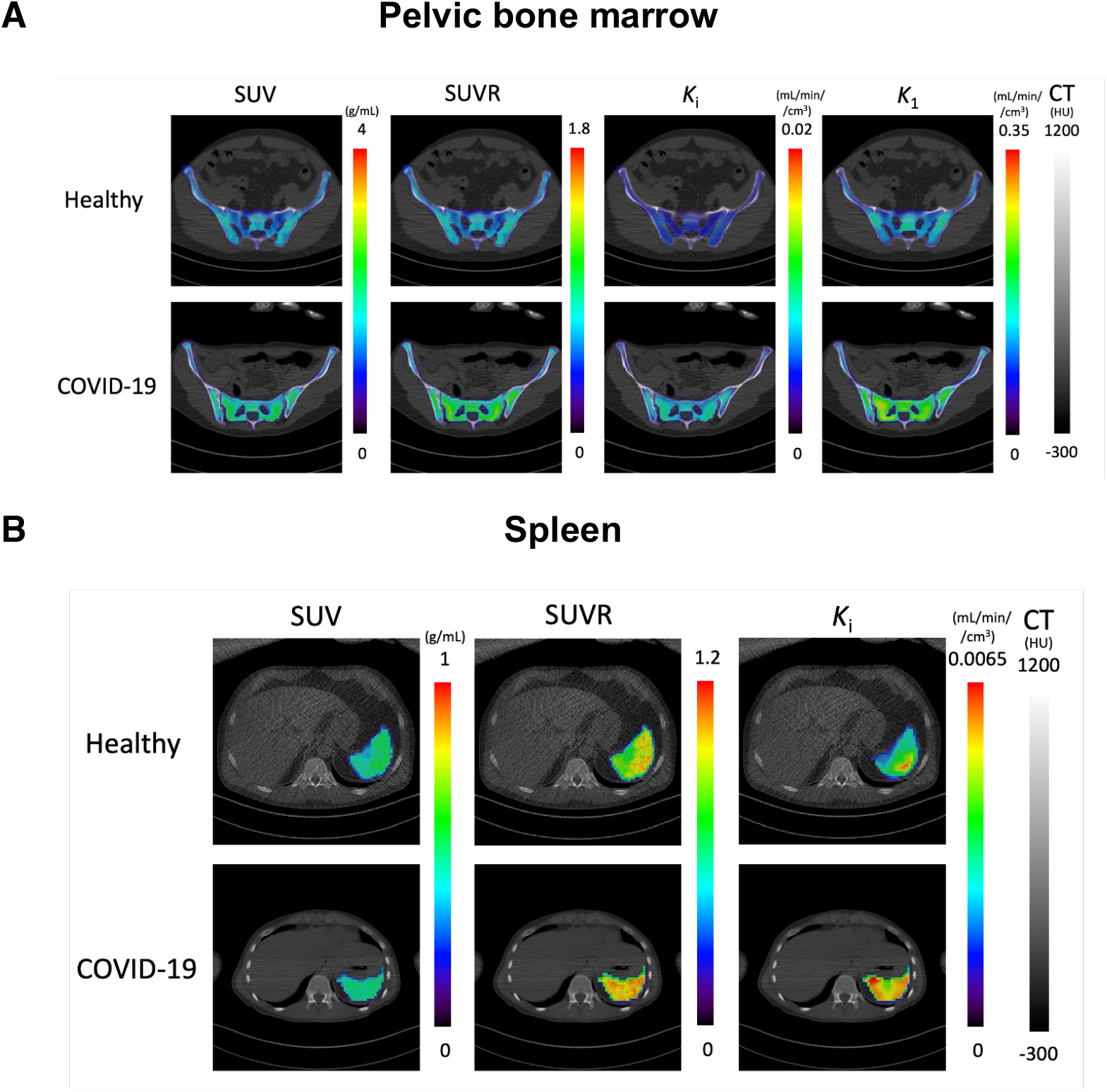
Parametric images of example healthy subjects and COVID-19 subjects. (A) Pelvic bone-marrow images of ^18^F-FDG SUV, SUVR, and parametric images of ^18^F-FDG net influx rate *K*_i_, and blood-to-tissue ^18^F-FDG delivery rate *K*_1_. (B) Spleen ^18^F-FDG SUV, SUVR, and parametric images of ^18^F-FDG net influx rate *K*_i_. The masked PET images are overlaid on the CT images.

## GRAPHICAL ABSTRACT

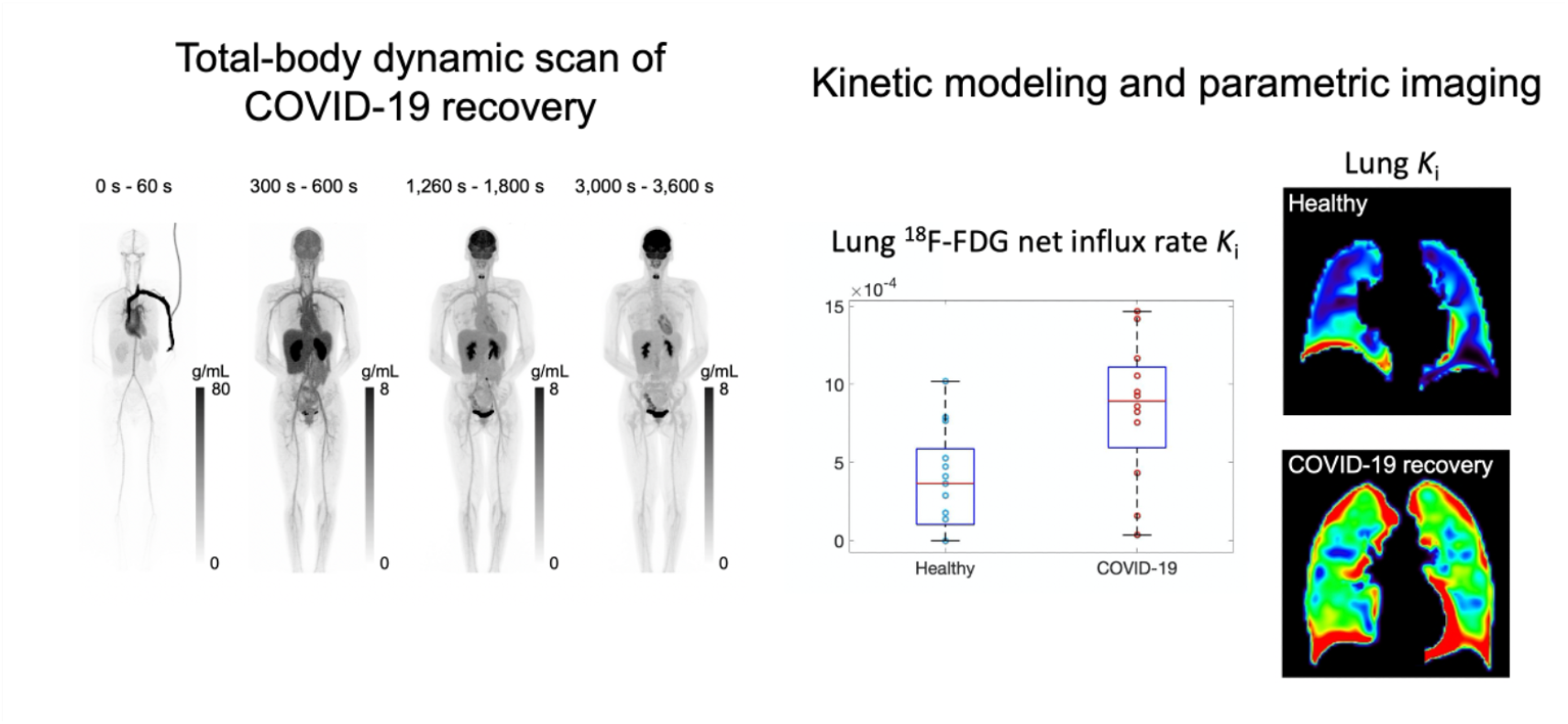

---

**SUPPLEMENTAL TABLE 1.** ROI Placement in Different Organs/Tissues for Kinetic Modelling

| Organ/tissue | ROI placement |
| --- | --- |
| Lung | Five same-sized spherical ROIs were placed in each of the five lung lobes. Large vessel structures and lung boundary were avoided to minimize the motion effect. The five lobe ROI TACs were extracted and averaged to acquire a global lung TAC. |
| Myocardium | A 3D free-hand ROI were placed in the myocardium according to both the late frames (45 min - 60 min) and the early frames (0 - 10 min) of the dynamic PET image to minimize the motion and the spill-over effects. |
| Liver | An ellipsoid ROI was placed in the liver. |
| Spleen | An ellipsoid ROI was placed carefully in the spleen to diminish the motion effect from the lung. |
| Spine bone marrow | Ten same-sized cylinder ROIs were placed in the bone marrow of ten spine sections (thoracic T8 - T12, and lumbar L1 - L5). The extracted ten TACs were averaged to acquire a global spine bone TAC. Both PET and CT images were referred. |
| Pelvic bone marrow | Six ellipsoid ROIs were placed in the pelvic bone marrow, three on the left and three on the right according to both the PET images and CT images. |
| Thigh muscle | An ellipsoid ROI was placed in the quadriceps femoris muscle of the right thigh and large blood vessels were avoided. |
| Gray matter | An isocontour ROI was placed in the gray matter according to the late phase (45 min - 60 min) PET image. |
| White matter | An ellipsoid ROI was placed in the white matter according to the PET image. |
| Brainstem | An ellipsoid ROI was placed in the brain stem according to the PET image. |
| Cerebellum | An ellipsoid ROI was placed in the cerebellum according to the PET image |
| Ascending Aorta | A 3D free-hand ROI was placed according to both the late frames (45 min - 60 min) and the early frames (0 - 10 min) of the dynamic PET images. |
| Right Ventricle | An ellipsoid ROI was placed according to both the late-frame (45 min - 60 min) and the early-frame (0 - 10 min) dynamic PET images. |

**SUPPLEMENTAL TABLE 2.** Information of individual subjects in the healthy controls and recovering COVID-19 subjects

| Subject index | Age (years) | Gender | Weight (kg) | BMI (kg/m<sup>2</sup>) | Dose (MBq) | Blood sugar level (mg/dL) | Fasting time before PET scan (hours) | COVID-19 vaccination |
| --- | --- | --- | --- | --- | --- | --- | --- | --- |
| H01 | 76-80 | Male | 71 | 24 | 349 | 101 | 11 | No |
| H02 | 51-55 | Female | 87 | 33 | 389 | 101 | 11 | No |
| H03 | 26-30 | Male | 112 | 34 | 387 | 77 | 6 | No |
| H04 | 46-50 | Male | 74 | 27 | 372 | 94 | 12 | No |
| H05 | 51-55 | Female | 67 | 24 | 348 | 93 | 12 | No |
| H06 | 61-65 | Male | 88 | 29 | 374 | 92 | 12 | No |
| H07 | 61-65 | Male | 80 | 24 | 376 | 79 | 12 | No |
| H08 | 46-50 | Male | 109 | 34 | 370 | 116 | 12 | No |
| H09 | 41-45 | Female | 53 | 19 | 389 | 78 | 11 | No |
| H10 | 51-55 | Female | 99 | 36 | 337 | 96 | 12 | No |
| H11 | 26-30 | Female | 81 | 30 | 370 | 100 | 12 | No |
| H12 | 26-30 | Female | 58 | 20 | 379 | 79 | 12 | No |
| H13 | 51-55 | Female | 89 | 35 | 390 | 91 | 10 | No |
| C01 | 31-35 | Female | 131 | 44 | 303 | 96 | 6 | No |
| C02 | 51-55 | Female | 106 | 39 | 309 | 86 | 8 | No |
| C03 | 41-45 | Female | 54 | 20 | 305 | 74 | 10 | Yes |
| C04 | 46-50 | Female | 72 | 29 | 292 | 83 | 12 | Yes |
| C05 | 36-40 | Male | 87 | 25 | 285 | 86 | 10 | No |
| C06 | 36-40 | Female | 69 | 25 | 298 | 81 | 12 | No |
| C07 | 51-55 | Male | 87 | 28 | 309 | 93 | 12 | Yes |
| C08 | 21-25 | Female | 59 | 19 | 275 | 74 | 12 | No |
| C09 | 46-50 | Female | 74 | 30 | 285 | 84 | 7 | Yes |
| C10 | 46-50 | Male | 57 | 19 | 244 | 93 | 12 | Yes |
| C11 | 26-30 | Female | 120 | 43 | 292 | 82 | 10 | Yes |
| C12 | 41-45 | Female | 89 | 31 | 290 | 93 | 12 | Yes |

Healthy controls are indexed as H01-H13, and COVID-19 recovering subjects are indexed as C01-C12.

